# Targeted proteomics upon Tofersen identifies candidate response markers for SOD1-linked amyotrophic lateral sclerosis

**DOI:** 10.1101/2024.04.22.24306165

**Authors:** Christina Steffke, Karthik Baskar, Maximilian Wiesenfarth, Johannes Dorst, Joachim Schuster, Florian Schöberl, Peter Reilich, Martin Regensburger, Alexander German, René Günther, Maximilian Vidovic, Susanne Petri, Jochen H. Weishaupt, Thomas Meyer, Tim Hagenacker, Julian Grosskreutz, Ute Weyen, Patrick Weydt, Thomas Haarmeier, Paul Lingor, Joachim Wolf, Andreas Hermann, Johannes Prudlo, Kornelia Günther, Antje Knehr, Zeynep Elmas, Özlem Parlak, Zeljko Uzelac, Simon Witzel, Wolfgang Philipp Ruf, Hayrettin Tumani, Albert C. Ludolph, Axel Freischmidt, Patrick Oeckl, Ritchie Ho, David Brenner, Alberto Catanese

## Abstract

Tofersen is the first effective and approved therapy for familial ALS caused by pathogenic variants in the *SOD1* gene. Following treatment with tofersen, neurofilaments in patients CSF and serum display a faster response than clinical parameters, underlining their importance as a biomarker for treatment response in clinical trials. This evidence led us to hypothesize that this novel treatment might represent an opportunity to identify additional therapy-responsive biomarkers for ALS. We chose the commercial NUcleic acid Linked Immuno-Sandwich Assay (NULISA™), to investigate a predefined panel of 120 neural, glial and inflammatory markers in CSF and serum samples longitudinally collected from SOD1-ALS patients at baseline and three months after tofersen treatment. We identified a set of proteins (beyond pNfH and NfL) whose levels differed between SOD1-ALS and the matched control group and that were responsive to treatment with tofersen, including Aβ42, NPY and UCHL1. Even though our results warrant validation in larger cohorts and longer follow-up time, they may pave the way for a panel of responsive proteins solidifying biomarker endpoints in clinical trials.

## Introduction

The VALOR trial and its open label extension (OLE) investigated the efficacy of the antisense oligonucleotide (ASO) tofersen for treating SOD1-mutant cases of ALS (SOD1-ALS) [1] and highlighted the great potential of surrogate and prediction markers for the clinical response to an anti-neurodegenerative therapy. The levels of the axonal marker neurofilament light chain (NfL) in serum and cerebrospinal fluid (CSF) indeed indicated a treatment response earlier than clinical functional scores [1], what we recently replicated under real-world conditions in a German cohort of SOD1-ALS patients treated with tofersen under the early access program (EAP) [2]. This observation encourages the use of neurofilaments as the primary endpoint for future clinical proof-of-concept trials in ALS [3-5]. However, since neurofilament levels potentially can be influenced by the drug candidate through other means than slowing down neurodegeneration (e.g., impact on its posttranslational modification, phosphorylation, turnover, or expression of *NEFH/NEFL*), a panel of response markers would be preferable for the sake of robustness of a biomarker-based primary endpoint. We hypothesized that tofersen as a drug with proven clinical efficacy, large effect size and immediate impact on validated neurodegeneration markers would offer, for the first time in the context of ALS, the opportunity to assess the therapy-responsiveness of other proteins. To this end, we leveraged the highly sensitive NUcleic acid Linked Immuno-Sandwich Assay (NULISA™) [6] to analyze a predefined panel of 120 targets in serum and CSF samples of nine SOD1-ALS patients treated with tofersen and nine matched control probands.

## Methods

Additional and detailed methods are described in the online Supplementary material.

### Participants

Nine SOD1-ALS patients were selected within the cohort previously published by Wiesenfarth et al. [2]. Individuals of the healthy control (HC) group suffered from non-neurodegenerative minor conditions including tension headache, idiopathic intracranial hypertension, and idiopathic facial nerve paralysis.

### NULISAseq assay

The NULISAseq CNS Disease Panel 120 was performed as previously described [6] (detailed information on the assay and data analysis can be found in the Supplementary Methods).

### Ethics

Biosampling of serum and CSF was conducted via participation in the MND-NET cohort study, for which informed consent was obtained. The study was approved by the local ethics committee of Ulm University (application number 19/12).

### Data availability

The raw NULISAseq data are available on request.

## Results

CSF and serum sample pairs were longitudinally collected from the same SOD1-ALS patients at baseline and at three months after tofersen treatment initiation, as well as from matched controls with minor neurological conditions (**Fig. 1A, Supplementary Table 1**). To specifically focus on treatment response, we selected nine patients from the German tofersen EAP who showed a clear drop in levels of neurofilament light (NfL) and heavy chain (NfH) after three months of treatment [2] (**Supplementary Table 1**). We focused on the early three-month time point to highlight immediate effect at the proteome level and minimise effects linked to disease progression.

**Figure 1.**
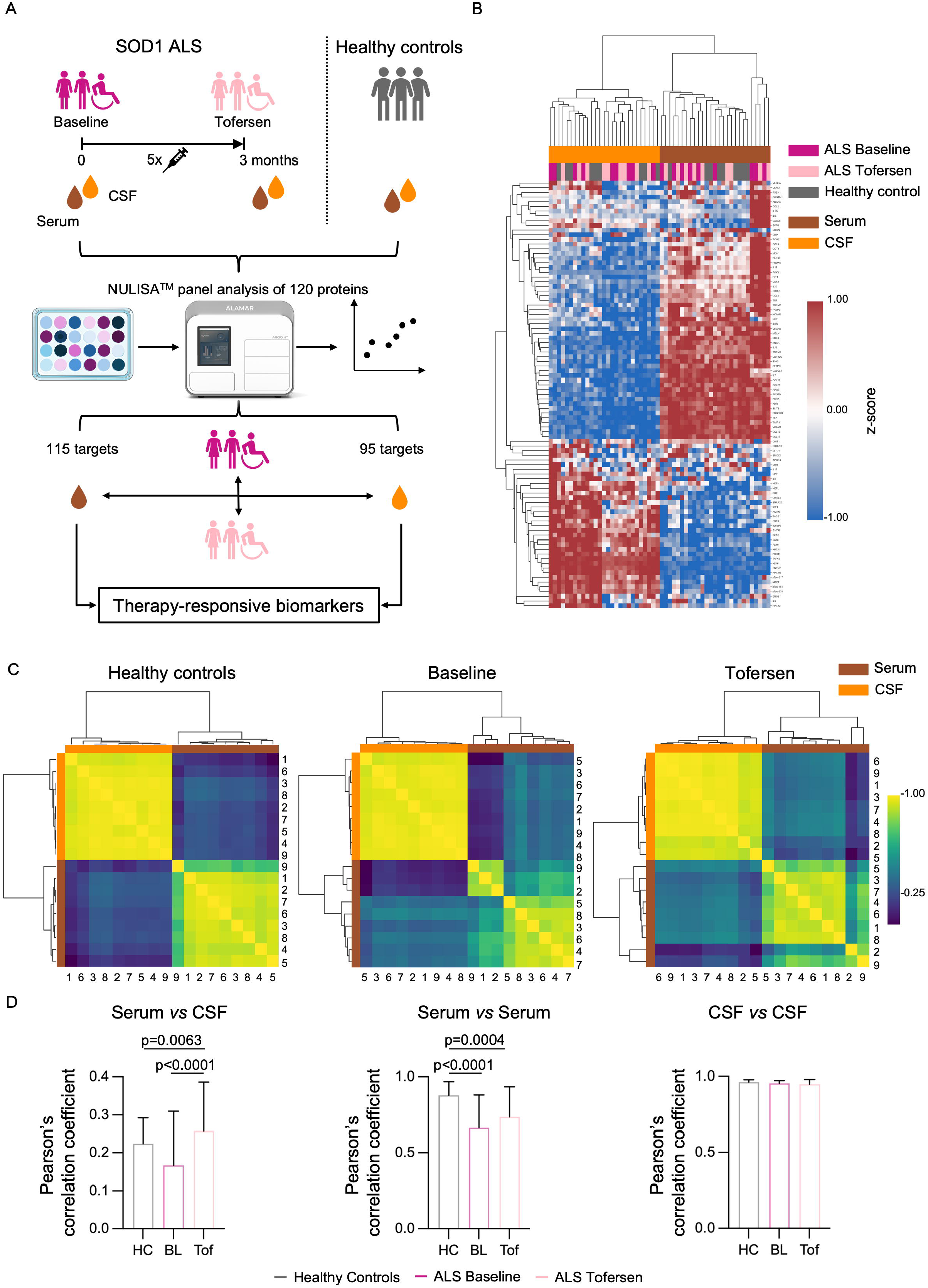
Serum and CSF show differential effects by tofersen. (A) Schematic representation of the study design: longitudinal collection of CSF and serum sample pairs from SOD1-ALS patients was performed at baseline and at 3 months after tofersen treatment initiation, as well as from age- and gender-matched controls with minor neurological conditions. Both biofluids were then analyzed with a NULISA™ panel platform including 120 proteins. (B) A clear separation of both biofluids is displayed after unbiased hierarchical clustering performed with the 91 proteins commonly detectable. (C) Healthy controls show a higher degree of homogeneity within serum and CSF targeted proteome than SOD1-ALS patients at baseline which was partially normalized upon tofersen treatment, accentuated by patient-centric correlation analysis. Numbers refer to patient numbers as listed in Supplementary Table 1. (D) Pearson’s correlation coefficients reveal increased heterogeneity in ALS-baseline samples compared to healthy controls (serum vs. CSF), which is normalized upon tofersen treatment. The highest contribution to this underlying heterogeneity rises from serum samples, while CSF shows comparable homogeneity across groups. Data are displayed as mean value ± SD (one-way ANOVA with post-hoc Tukey test). Statistical significance was set at p< 0.05, exact p-values are displayed.

We identified 115 proteins for serum and 95 for CSF samples that passed quality control and were selected for further analysis (**Supplementary Fig. 1A**). Unbiased hierarchical clustering performed with the 91 proteins commonly detectable showed a clear separation of the two body fluids but could not cluster the samples according to the three groups (healthy controls, SOD1-ALS baseline and SOD1-ALS tofersen) (**Fig. 1B**). We then performed principal component analysis (PCA) to explore the multiple dimensions (i.e., genotype, treatment and biofluid) of our data (**Supplementary Fig. 1B**). Even when we considered the top six and most meaningful principal components (PCs) of this mixed dataset (**Supplementary Fig. 1C**) we could not effectively separate the three conditions (**Supplementary Fig. 1D**), indicating that the samples from the two biofluids might be influenced by different sources of variability. Accordingly, patient-centric correlation analysis highlighted a higher degree of heterogeneity within the serum and CSF targeted proteome of the SOD1-ALS patients at baseline than in the healthy controls. While a higher degree of similarity was observed in CSF, serum samples displayed the highest inter-patient heterogeneity, which was partially normalized upon tofersen treatment (**Fig. 1C-D**). Thus, we proceeded to analyze both biofluids separately.

We first focused on serum, in which the SOD1-ALS samples displayed the highest degree of variability. By comparing the protein panel of SOD1-ALS patients at baseline to controls, we identified NfL, the Tau isoforms pTau-231, pTau-181 and total Tau (MAPT) as being significantly up-regulated after correction for multiple comparisons. When looking at the unadjusted p-value, 26 molecules were significantly altered in the ALS group: IL-15, IL-16 and Aβ40 were downregulated while the levels of the aforementioned markers, other interleukins and several cytokines were higher in disease samples (**Supplementary Fig. 2A-C**). This confirmed that the NULISA™ technology can identify a SOD1-ALS fingerprint in serum. Nevertheless, despite SOD1-ALS samples could be effectively separated from controls by PCA when considering PC2 and PC3, the tofersen samples remained close to their baseline counterparts (**Fig. 2A-B**). Accordingly, we did not observe any significant correlation (**Fig. 2C**; Spearman Correlation: 0.02) between the effect of the ASO on the serum panel targets and on disease progression. Since additional deeper exploration of the different PCs also did not reveal any significant effect linked to the treatment (**Supplementary Fig. 3A-B**), we looked at the combined PC-loadings of PC2 and PC3. This approach highlighted APOE4, NfH and NfL as the proteins mostly driving the shift from untreated to treated state (**Fig. 2D; Supplementary Fig. 4**). In agreement with these findings, paired comparison (without correction for multiple comparisons) of the untreated and treated SOD1-ALS samples confirmed a significant reduction of NfH and NfL [1,2] . Moreover, we detected a significant increase of GFAP, IL-9, SMOC1, NPTX2, Aβ40, IL-15, TAFA5, NPY, and VCAM1 upon tofersen treatment (**Fig. 2E; Supplementary Fig. 5**). Apart from neurofilaments, only Aβ40 was inversely altered in untreated SOD1-ALS vs. controls, indicating only these markers were reverted by tofersen in serum (**Table 1**).

**Figure 2.**
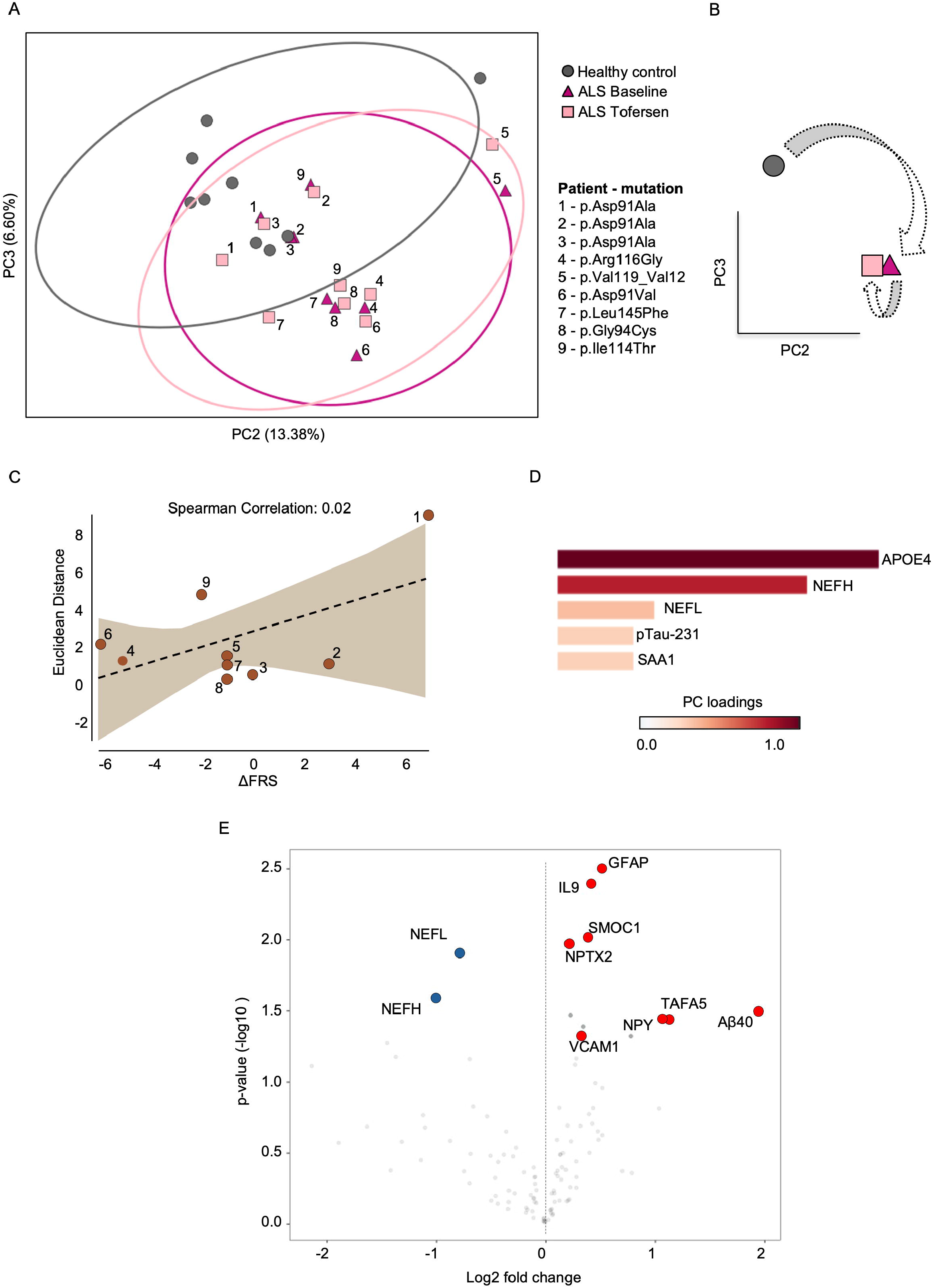
Tofersen causes only minor changes to the panel targets in the serum of SOD1-ALS patients after three months of treatment. (A) PC2 and PC3 distinguish SOD1-ALS from healthy controls (HC) but cannot separate the patients on the basis of ASO treatment, as highlighted in (B) centroid representation of PC2 and PC3. (C) The effect of tofersen treatment on the serum targeted proteome (analyzed as Euclidean distance of the paired ALS patients at baseline and after tofersen treatment along PC2 and PC3) does not significantly correlate with clinical progression (calculated as the difference between the ALS-FRS score after 3 months of treatment and at baseline). Euclidean distance was calculated as follows: using the x- and y-coordinates (x_1_y_1_ at baseline, x_2_y_2_ after 3 months of tofersen) for each patient: 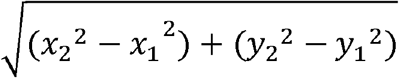. Spearman’s correlation was performed. (D) PC-loadings of PC2 and PC3 combined reveal APOE4 and neurofilaments as driving markers from untreated to treated state. (E) Volcano plot displays significantly altered panel targets between baseline and tofersen-treated SOD1-ALS samples. Significance was calculated using the linear mixed effect model. Statistical significance was set at p< 0.05.

**Figure 3.**
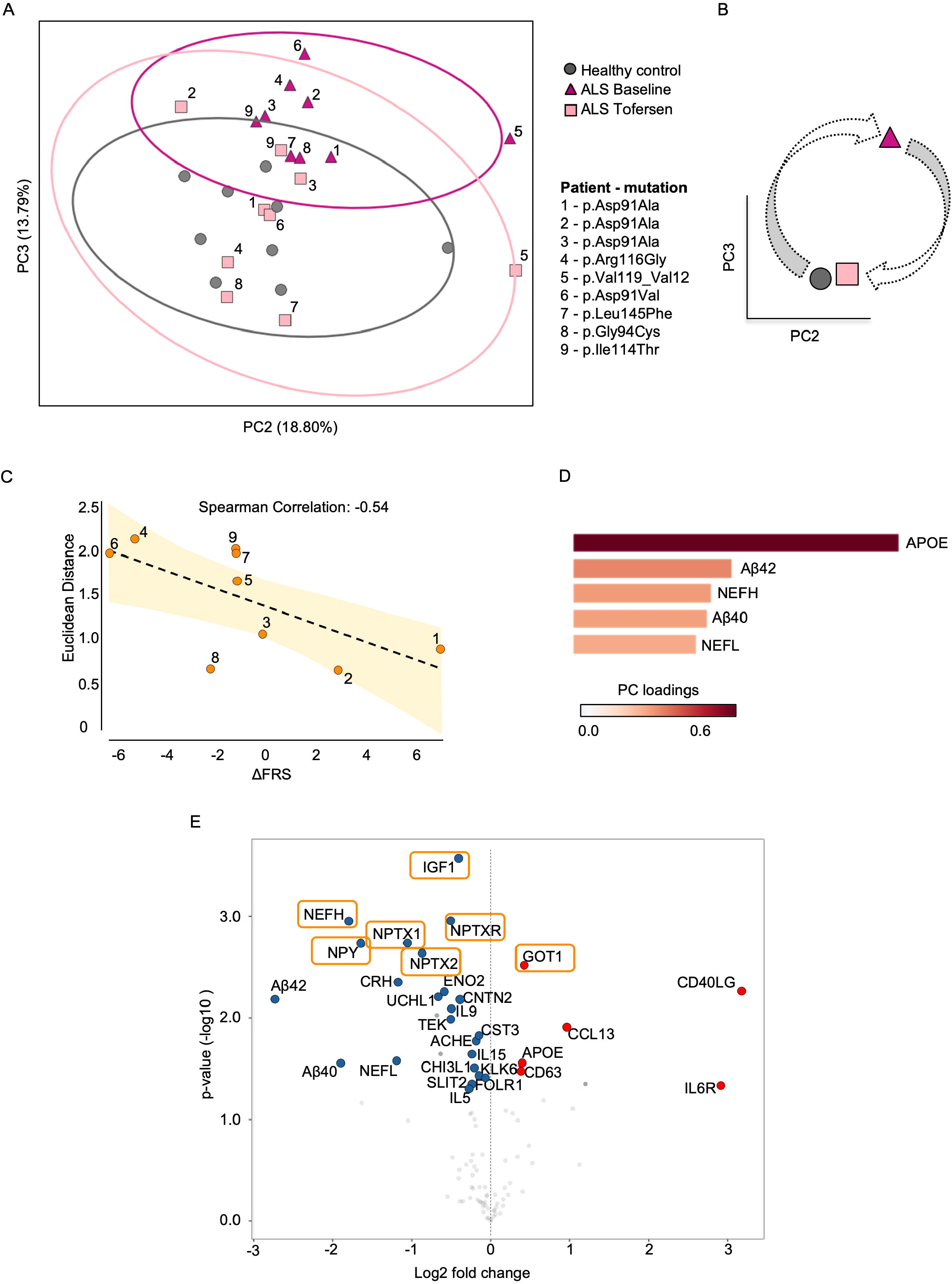
CSF displays a proteomic fingerprint of neuroprotection upon tofersen treatment. (A) PC2 and PC3 display a clear transition from tofersen-treated patients close to the healthy controls, underlined by (B) Centroid representation of PC2 and PC3. (C) Alterations of panel targets in CSF inversely correlate (determined through Spearman’s correlation) with clinical progression (Euclidean distance and progression rate were calculated as described above). (D) APOE4, Aβ42, NfH, Aβ40 and NfL have the highest load in of PC2 and PC3 combined. (E) Volcano plot displays significantly altered panel targets between baseline and tofersen-treated SOD1-ALS samples. Significance was calculated using the linear mixed effect model. Orange boxes highlight significant markers according to adjusted p-value. Statistical significance was set at p< 0.05.

**Table 1.**
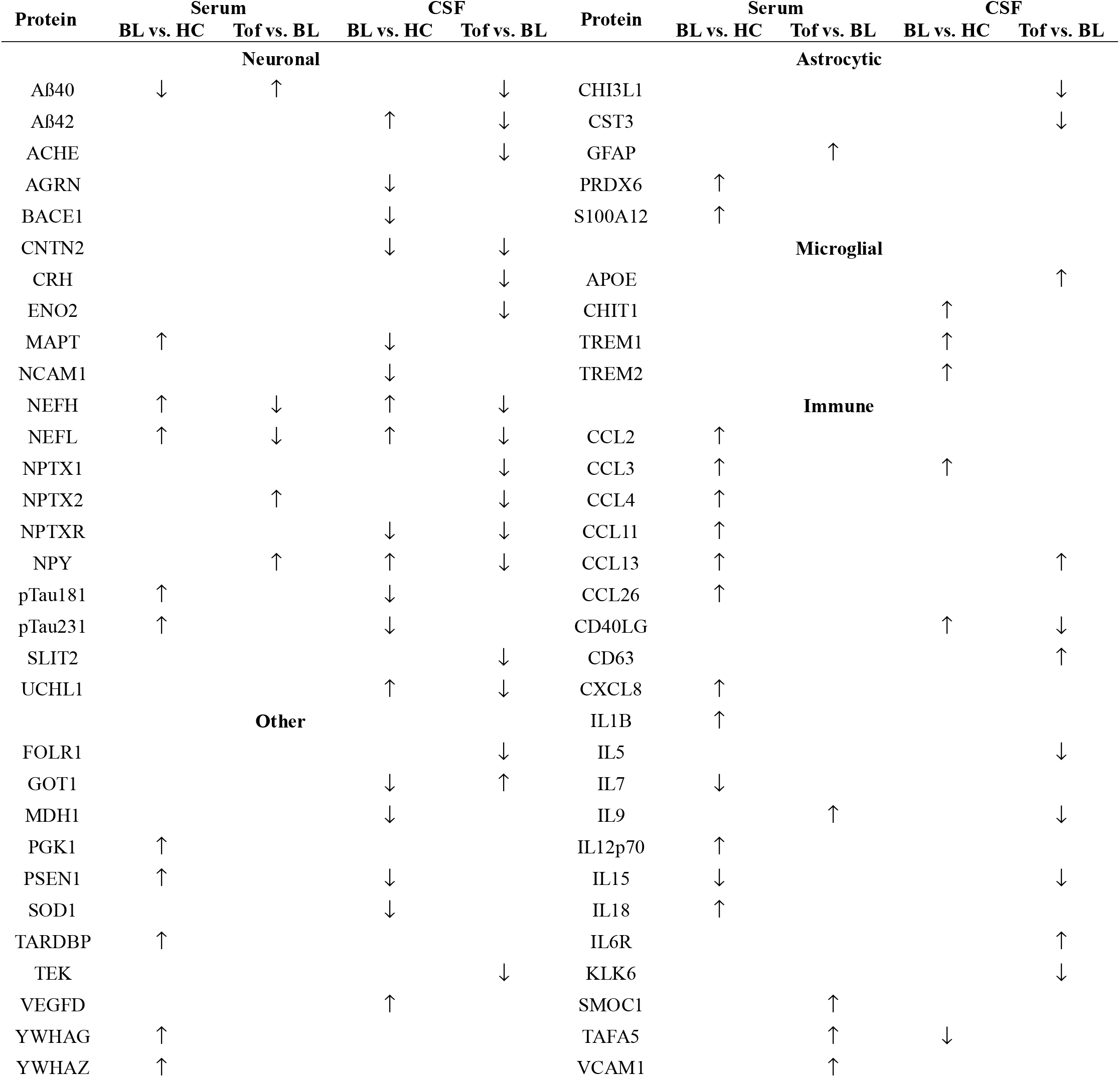
Significantly altered panel targets in serum and CSF between SOD1-ALS baseline (BL) and healthy controls (HC) and between tofersen-treated (Tof) and non-treated state (BL). Arrows indicate down-or up-regulation.

We then investigated the effect of tofersen treatment in the CSF. First, we compared untreated SOD1-ALS to controls and found that the levels of several established ALS biomarkers such as NfH, NfL, UCHL1 and CHIT1 were increased in patients (**Supplementary Fig. 6A-B**). The proteins NCAM1, NPTXR, GOT1, PSEN1, SOD1 and different Tau forms were significantly decreased in SOD1-ALS compared to controls (**Supplementary Fig. 6C**). According to these alterations in the CSF of SOD1-ALS patients, PC1 could clearly separate the disease status from controls but, when used in combination with PC2, could not distinguish between untreated and tofersen samples. We then combined PC1 and PC3 highlighting a solid shift of the tofersen towards the control group along PC3, even though the effect of PC1 was still enough to separate the SOD1-ALS patients treated with tofersen from the healthy control group (**Supplementary Fig. 7**). Thus, we reasoned that PC2-PC3 might represent the best combination to identify a neuroprotective effect driven by tofersen. In fact, by using these two PCs we could observe a clear shift of the ALS samples upon treatment, which moved back close to the healthy control group (**Fig. 3A-B**). This indicated that the proteins responsible for this shift might represent the fingerprint for a neuroprotective effect in SOD1-ALS. We indeed observed a high, inverse correlation (Spearman value: -0.54; **Fig. 3C**) between the changes in the protein panel and disease progression. Interestingly, the lowest impact on the protein levels was detected in two slow-progressor patients (# 1 and 2; both p.Asp91Ala carriers) whose ALS functional rating scale revised (ALSFRS-R) delta scores even ameliorated after three months of treatment (**Supplementary Table 1**). This suggests that, in this short period, the ASO might have exerted more profound molecular effects on patients with faster disease progression. Therefore, we aimed at identifying the panel targets underlying the effect of the ASO and looked at the PC-loadings in an analogous manner as for the serum samples. We identified APOE4, Aβ42, NfH, Aβ40, NfL and NPY as the proteins with the highest significance in PC2 and PC3 (**Fig. 3D; Supplementary Fig. 8**). We then performed a paired comparison of CSF samples from SOD1-ALS patients at baseline with 3 months of tofersen and found 23 and 6 proteins being significantly down-or up-regulated, respectively. Specifically, the levels of ACHE, Aβ40, Aβ42, CHI3L, CNTN2, CRH, CST3, ENO2, FOLR1, IGF1, IL-5, IL-9, IL-15, KLK6, NEFH, NEFL, NPTX1, NPTX2, NPTXR, NPY, SLIT2, TEK and UCHL1 were reduced by the treatment, while APOE, CCL13, CD40LG, CD63 and GOT1 were increased (**Fig. 3E; Supplementary Fig. 9)**. Of note, GOT1, NPY, Aβ42, CD40LG, and UCHL1 were inversely altered in untreated SOD1-ALS *versus* controls, indicating that these markers could be useful for both for diagnostic and therapeutic purposes (**Table 1**).

## Discussion

This study assessed the short-term effect of tofersen treatment on a targeted portion of the CSF and serum proteome from SOD1-ALS patients. We opted for the multiplexed NULISA™ platform [6], which allowed us to test the therapy-responsiveness of 120 apoptosis, neural, astrocytic, microglial and inflammatory markers with attomolar sensitivity at the same time. The reliability of this approach was confirmed by comparing SOD1-ALS patients before treatment to matched control probands, which highlighted altered levels of previously-described ALS biomarkers such as the neurofilaments, Tau-isoforms and molecules linked to immune and glial responses. Most notably, we found a fraction of markers, namely Aβ42, NfH, NfL, NPY,UCHL1 and GOT1, which not only discriminated SOD1-ALS from controls, but whose levels were also partially “corrected” by the ASO, thus potentially qualifying them as dual diagnostic and therapeutic markers for SOD1-ALS (and possibly also for ALS in general). Consistent with previous studies in sporadic ALS [7-10], the CSF levels of Aβ42 were increased in SOD1-ALS patients and normalised following treatment with tofersen. Since Aβ40 showed a similar trend, our results reinforce the previously reported interplay between SOD1 and Aβ peptides in ALS [11]. In fact, Aβ seems to accumulate in neurons, muscle/neuromuscular junctions and skin of ALS patients, possibly associated with oxidative stress [12-15]. In addition, both APP and BACE1 are upregulated in SOD1^G93A^ transgenic mice [16,17], while inhibition of APP processing proved neuroprotective in the same model [18]. Hypothetically, tofersen may either indirectly (e.g. by alleviating oxidative stress) reduce the expression of APP or impact the pathological cleavage of Aβ peptides. Both possibilities, which require further mechanistic investigations, may result in a lowered release of Aβ species in the extracellular environment and, consequently, into the CSF.

A similar principle might also apply to NPY, which is mainly expressed by interneurons, and whose increase in the CSF of SOD1-ALS patients might represent an endogenous, compensatory (synaptic) mechanism to counteract apoptosis and inflammation [19]. In fact, NPY expression was found to be higher in human post-mortem ALS motor cortex than in controls, and antagonism of the Y1 receptor is sufficient to rescue motor phenotypes in a murine model of ALS [20]. In this context, this data support the notion that re-establishing the excitation/inhibition balance might be an effective therapeutic intervention to contrast motor neuron degeneration. Indeed, also the neuronal pentraxins NPTX1 and NPTX2 and their receptor NPTXR also showed a robust class effect following treatment with tofersen, albeit they did not discriminate between untreated SOD1-ALS and controls. The neuropentraxins are candidate markers of synaptic dysfunction in cognitive disorders [21], and even if an interaction with SOD1 has not been described yet, they might also reflect a beneficial effect on synaptic homeostasis upon tofersen exposure. Accordingly, the well-established neural damage marker ENO2 (neuron-specific enolase), the axonal/synaptic markers Contactin-2 and AChE (acetylcholinesterase), as well as the apoptosis marker Annexin A5 were decreased after tofersen treatment while GOT1 (glutamate oxaloacetate transaminase 1), which acts as a scavenger of glutamate in the brain, was increased upon ASO therapy. The normalization of GOT1 levels, which are lower in ALS patients than controls, might be interpreted as “metabolic normalization” and hypothetically may increase the capacity to counteract glutamate excitotoxicity. Interestingly, combined treatment with recombinant GOT and its co-factor oxaloacetic acid improved motor neuron disease in an ALS rat model [22]. All these findings suggest that tofersen might exert neuroprotection at the cellular levels already after three months of treatment (if not earlier): as in the case of the neurofilaments, a pronounced extracellular release of UCHL1 is observed upon degeneration [23-25] and tofersen effectively reduced the levels of UCHL1 in CSF, and of NfH and NfL in both biofluids. In general, analysis of CSF yielded more robust results, likely due to its close contact to the central nervous system and its purer composition with less background noise factors. In contrast, the blood proteome might reflect a more diluted effect of tofersen. The serum samples showed indeed a higher degree of heterogeneity and several inflammatory and glial markers displayed a heterogenous behaviour in blood and CSF upon tofersen treatment (**Table 1**), possibly reflecting differential changes in cell types as well as inflammatory pathways. Of note, the well-established ALS microglial marker CHIT-1 was elevated in CSF of SOD1-ALS patients as previously described [26,27], but did not show a response to tofersen treatment. Some of the changes in glial and inflammatory markers may directly depend on SOD1 lowering, some may be drug-dependent but SOD1-unrelated, while others might occur secondary to alleviated neurodegeneration. Thus, a follow-up study including later time points is warranted to untangle the complex interplay between tofersen treatment and inflammatory and glial response.

Conclusively, our findings suggest that a panel of biomarkers may yield an increased informative value on whether and how robustly a drug influences different aspects of disease pathology (e.g., neurodegeneration, gliosis, inflammation). Even though analysis of larger cohorts and additional time points is required to understand how tofersen ultimately influences the disease in SOD1-ALS, we highlighted novel and essential features of neuroprotection achieved in ALS patients. Given validation and refinement, such a panel could represent a valuable readout for future clinical trials.

## Data Availability

All data produced in the present study are available upon reasonable request to the authortofersen under the early access program

## Acknowledgements

We thank patients and control probands for their donation of biosamples. We acknowledge the support of the coworkers of the Biomaterial repository of the Neurology Department of University Hospital of Ulm. This study was supported by a grant from Alamar Biosciences. The NULISAseq assay and analysis was performed at Alamar Biosciences in Fremont, California. We would like to extend our gratitude to the Technology Access Program team for their assistance in generating the data and collaborating on the analysis.

## Funding

This project was financially supported by the Deutsche Gesellschaft für Muskelkranke (DGM) project Ca2/1, the Deutsche Forschungsgemeinschaft (DFG; German Research Foundation) through SFB 1506 -Project A01 and the Alamar Neurology Grant to AC. AC and DB are also supported by the Frick Foundation for ALS research. AC receives further support from the Else Kröner-Fresenius Stiftung (project Nr. 2019_A111; L.SBN.0162) the Karin Christiane Conradi Foundation, the Deutsche Forschungsgemeinschaft (individual project CA 2915/4-1) and the Medical Scientist Program of the Ulm Medical Faculty. DB receives funding from the Else Kröner-Fresenius-Stiftung (Exzellenzstipendium, 2022_EKES.18) and the Deutsche Gesellschaft für Muskelkranke (DGM) (Br1/1). MR was funded by the BMBF (Clinician Scientist Programme ACS_iIMMUNE, 01EO2105). SP has received research support from NDR, the German Neuromuscular Society (DGM) and the German Israeli Foundation (GIF).

## Competing interests

SP has received speaker fees from Amylyx, Biogen, ITF-Pharma, Roche, and Zambon, and served on advisory boards for Amylyx, Biogen, Roche, Zambon, and ITF Pharma, and has participated as an investigator on clinical trials on ALS sponsored by Alexion Pharmaceuticals, AL-S Pharma, Amylyx, Apellis Pharmaceuticals, Biogen, Cytokinetics, Corcept Therapeutics, Ferrer, Sanofi, Orion Pharma, and Orphazyme. FS reports honoraria for advisory boards of Alnylam, Amylax and Alexion. ZU has received honoraria and grants from Biogen as a consultant. MV received travel expenses and non-financial support from ITF Pharma outside the submitted work. DB owns stocks from IONIS Pharmaceuticals. The other authors report no competing interests.

## Notes

### Clinical Trial

tofersen early access program

### Funding Statement

This study was funded by the Deutsche Gesellschaft fur Muskelkranke (DGM) project Ca2/1, the Deutsche Forschungsgemeinschaft (DFG; German Research Foundation) through SFB 1506 - Project A01 and the Alamar Neurology Grant to AC

