## Supplementary information for "Targeted proteomics upon Tofersen identifies candidate response markers for SOD1-linked amyotrophic lateral sclerosis"

**Running title:** New therapy-responsive markers for SOD1-ALS

**Keywords:** amyotrophic lateral sclerosis, tofersen, SOD1, ASO, serum, CSF

Supplementary material: this file contains 9 supplementary figures, 1 supplementary table and supplementary information regarding the methods used in the study

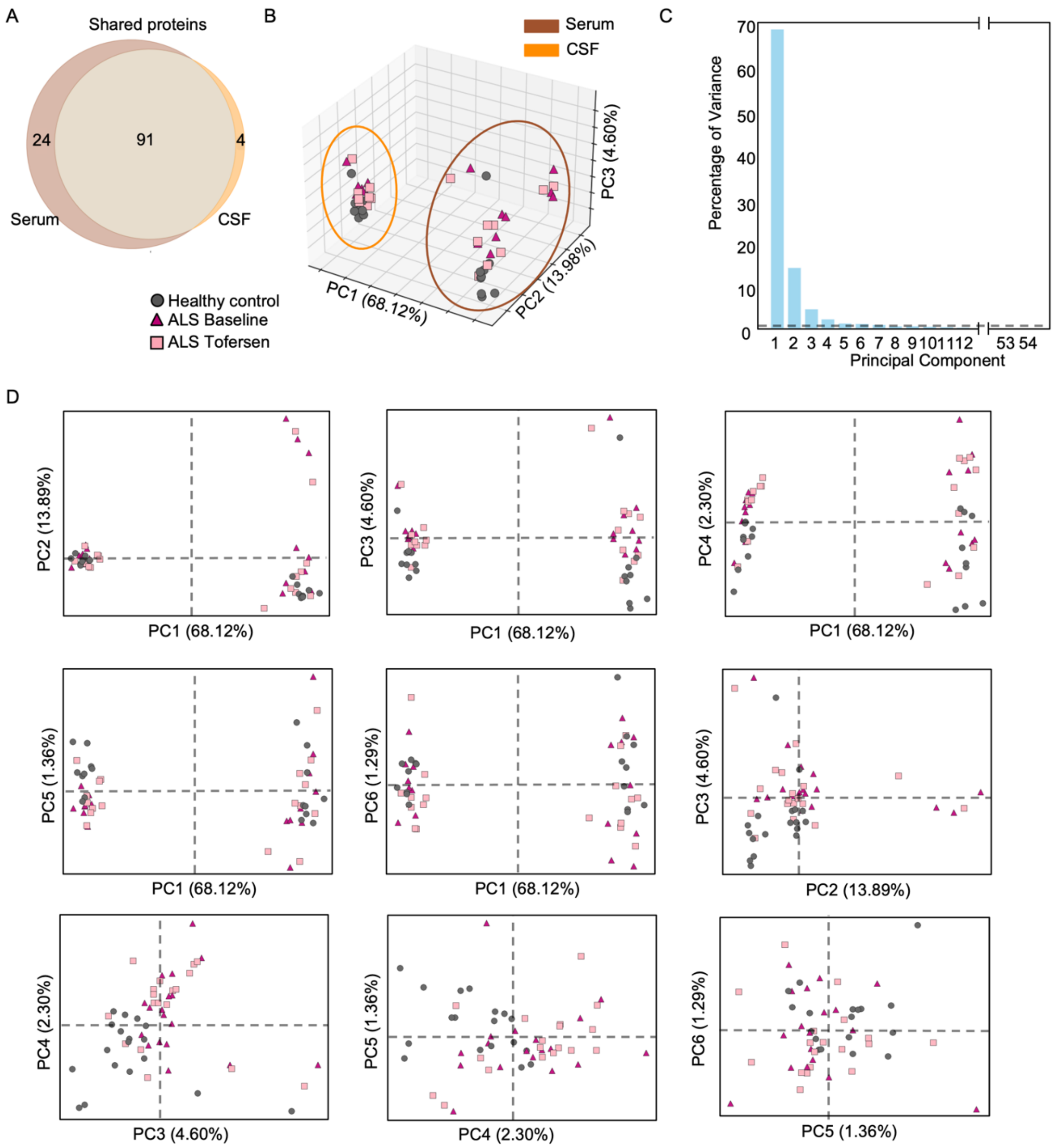


**Supplementary Fig. 1 Analysis of commonly detectable proteins in serum and CSF.**

(A) Venn diagram displays 91 commonly detectable proteins in both biofluids. (B) 3D-representation of PC1, PC2 and PC3 shows clear separation between serum and CSF. PC-analysis according to (C) percentage of variance (cut-off for principal components is depicted by the dotted line and was calculated as follows: 100/total number of PCs -1) of (D) 6 most meaningful PCs could not reveal any separation between the three sample conditions.

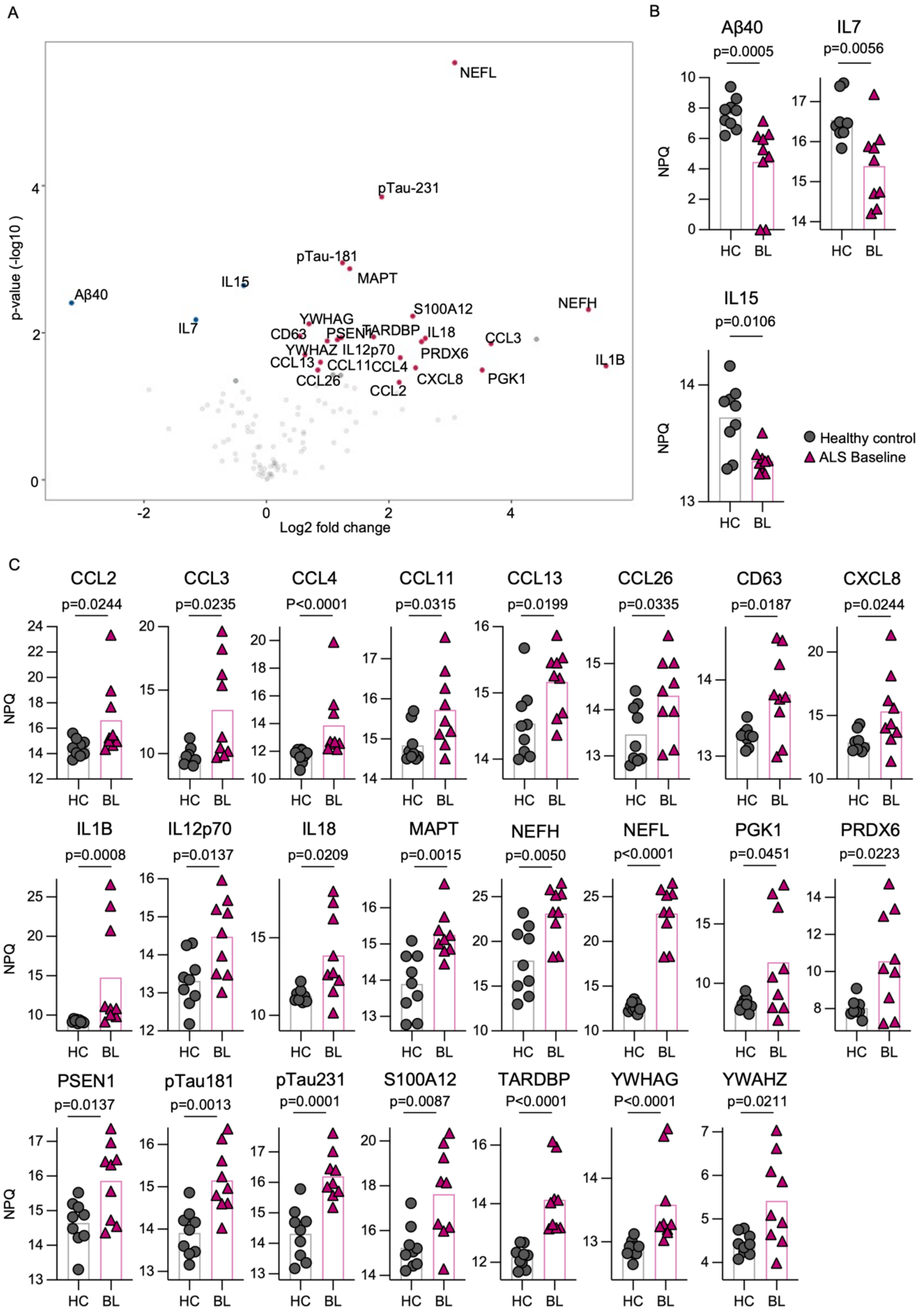


**Supplementary Fig. 2 Comparison between ALS-baseline and healthy controls in serum.** (A) Significantly altered proteins in a linear regression model displayed by volcano plot were further analysed by t-testing (Welch's t-test or Mann-Whitney test), revealing significantly (B)

downregulated and (C) upregulated proteins. Statistical significance was set at $p < 0.05$, exact p-values are displayed. Data are log2 transformed.

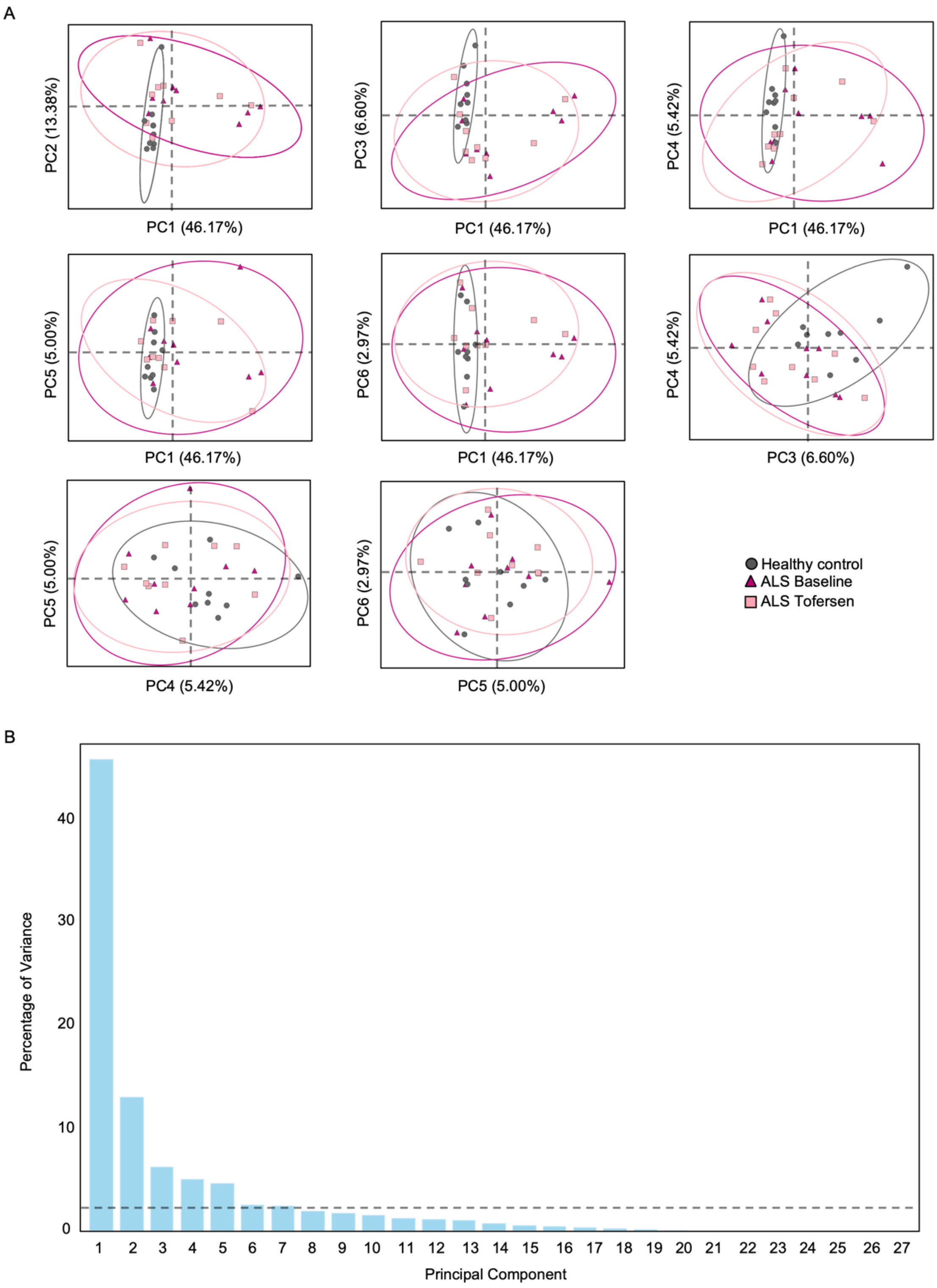


**Supplementary Fig. 3 Additional analysis in serum between all three conditions.**

(A) Further PC-analysis could not show major effect of tofersen on panel proteins. Meaningful PCs were selected according to (B) percentage of variance (cut-off for principal components is depicted by the dotted line and was calculated as follows: 100/total number of PCs -1).

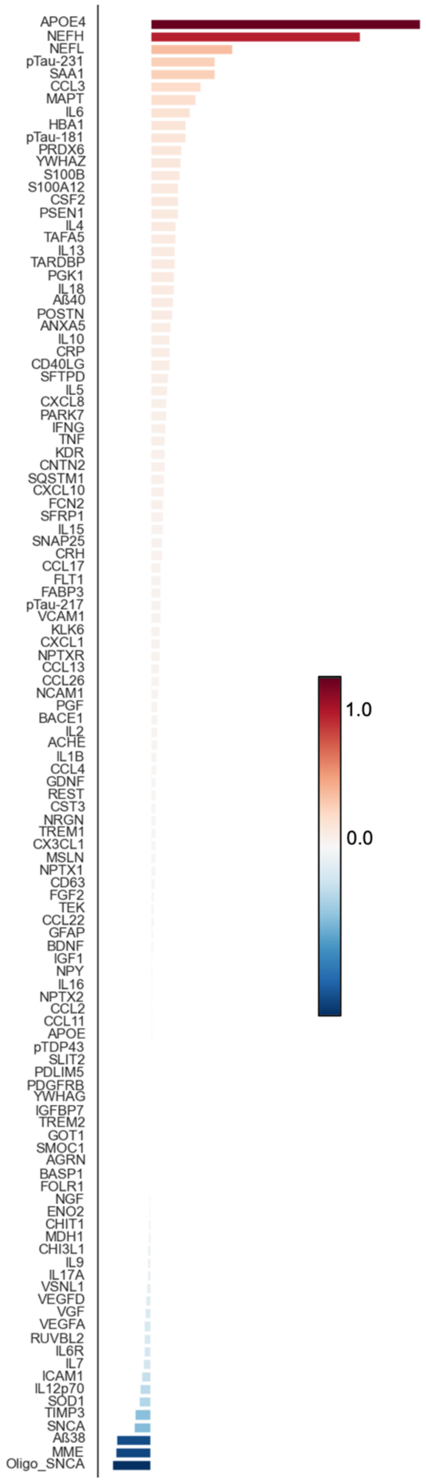


**Supplementary Fig. 4 Complete PC-loadings of PC2 and PC3 combined in serum.**

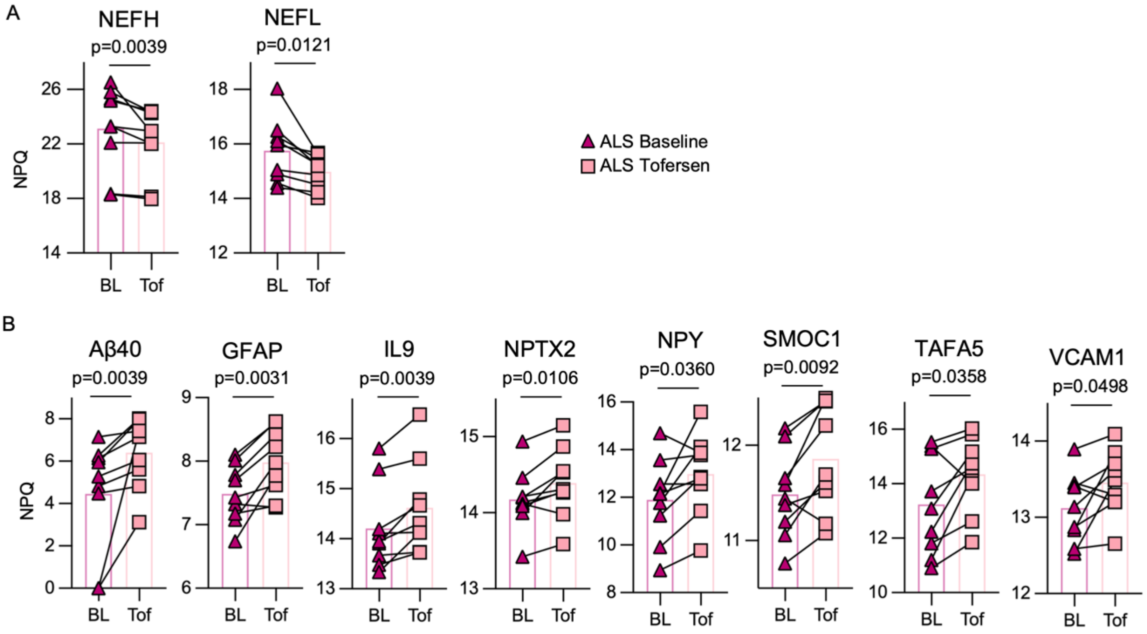


**Supplementary Fig. 5 Paired analysis of significantly changed proteins according to linear mixed effect model between tofersen and baseline condition in serum.**

Significantly (A) downregulated and (B) upregulated proteins. Paired t-test was applied. Statistical significance was set at $p < 0.05$, exact p-values are displayed. Data are log2 transformed.

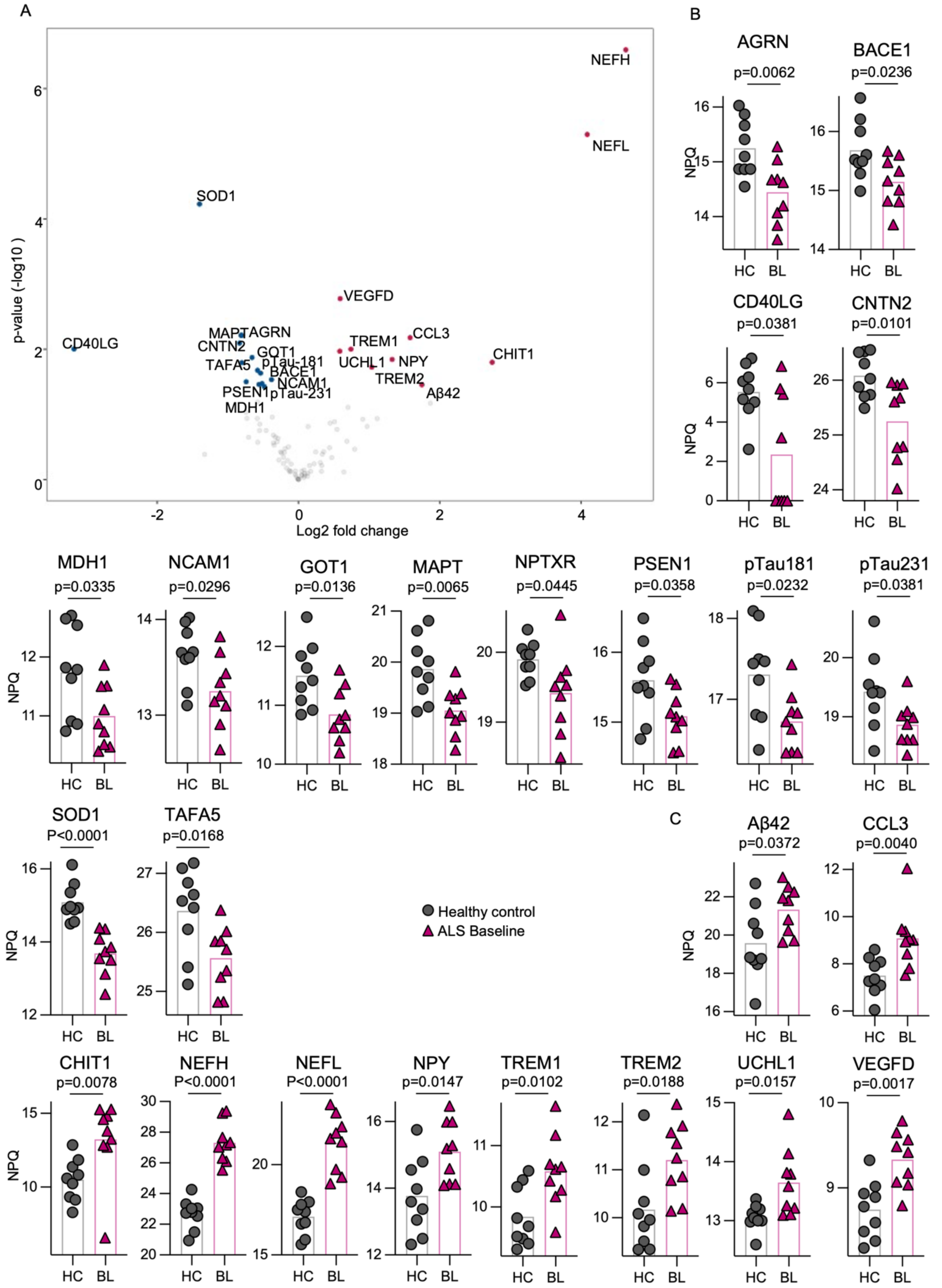


**Supplementary Fig. 6 Comparison between ALS-baseline and healthy controls in CSF.**
(A) Significantly altered proteins in a linear regression model displayed by volcano plot were further analysed by t-testing (Welch's t-test or Mann-Whitney test), revealing significantly (B) downregulated and (C) upregulated proteins. Statistical significance was set at $p < 0.05$, exact p-values are displayed. Data are log2 transformed

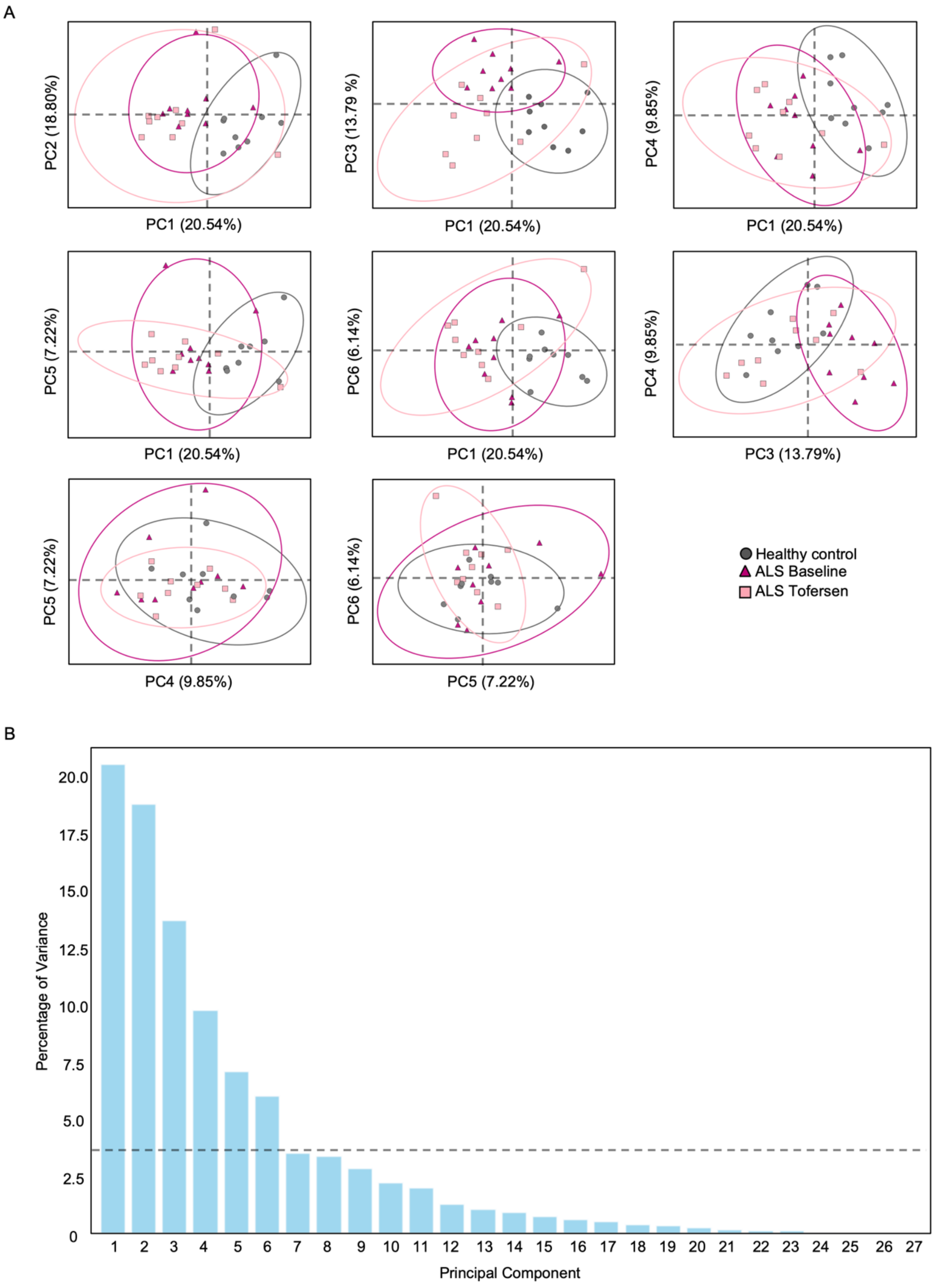


**Supplementary Fig. 7 Additional analysis in CSF between all three conditions.**

(A) Further PC-analysis could show separation between ALS and healthy conditions by PC1. Meaningful PCs were selected according to (B) percentage of variance (cut-off for principal components is depicted by the dotted line and was calculated as follows: 100/total number of PCs -1).

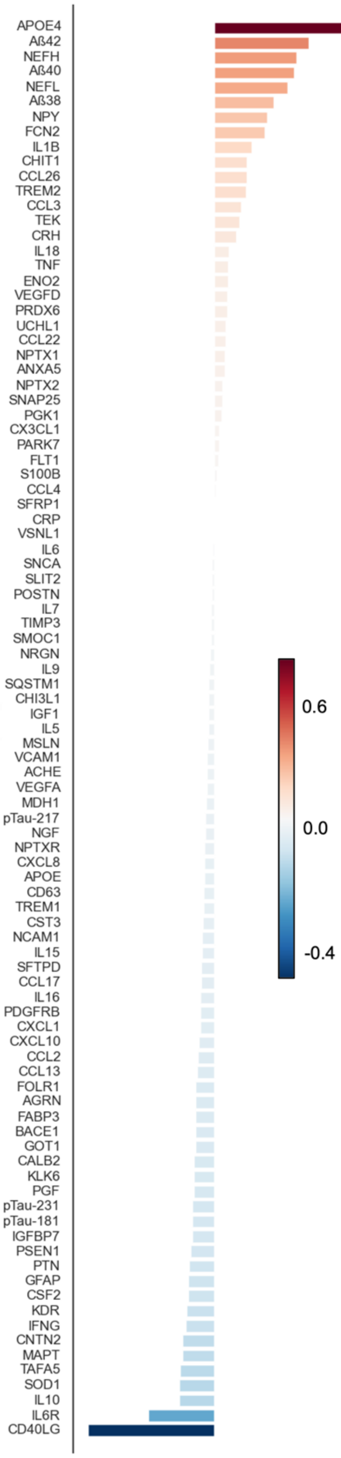


**Supplementary Fig. 8 Complete PC-loadings of PC2 and PC3 combined in CSF.**

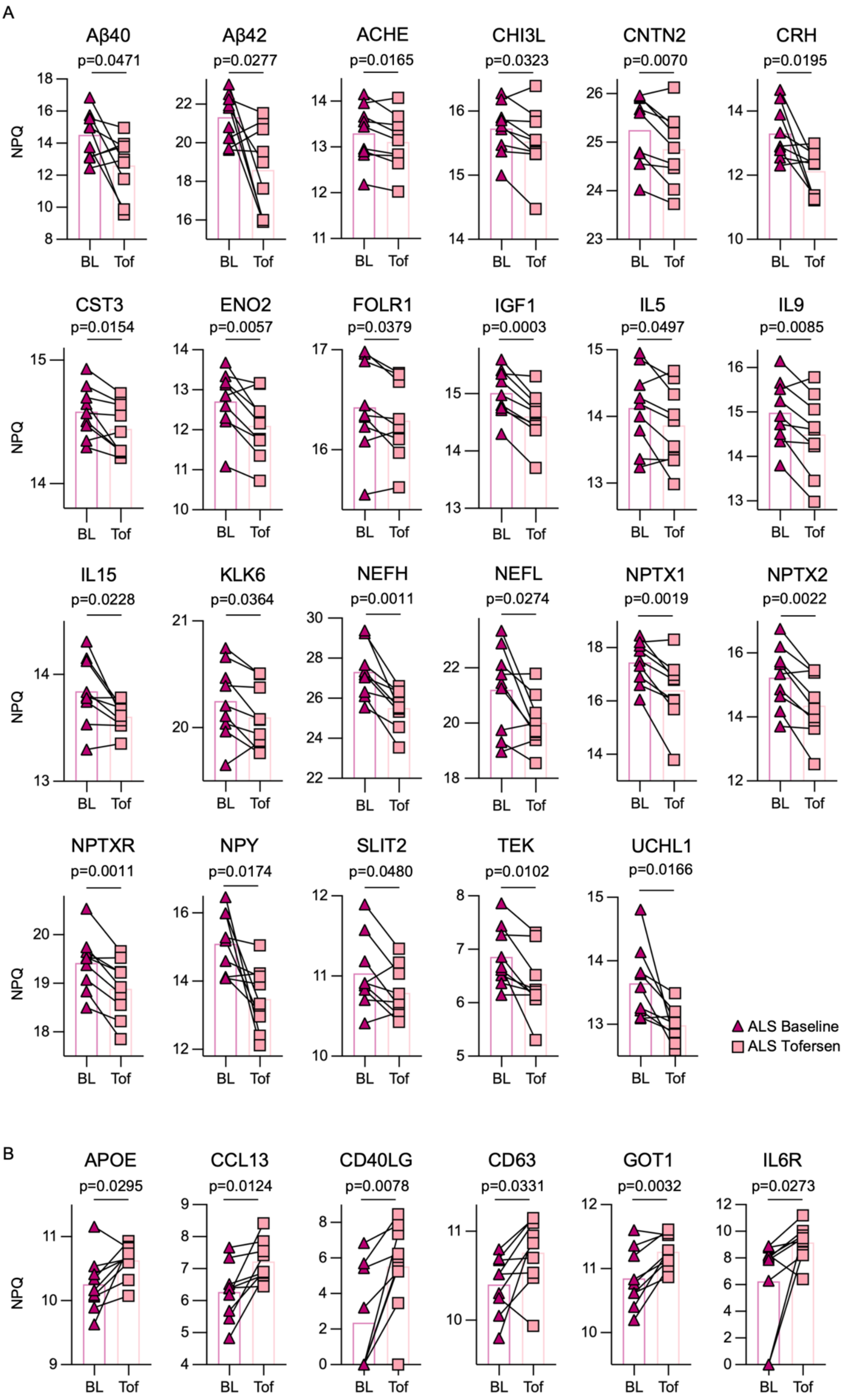


**Supplementary Fig. 9 Paired analysis of significantly changed proteins according to linear mixed effect model between tofersen and baseline condition in CSF.**

Significantly (A) downregulated and (B) upregulated proteins. Paired t-test was applied. Statistical significance was set at $p < 0.05$, exact p-values are displayed. Data are log2 transformed.

#### Supplementary Table 1 Clinical information of the participants included in the study

| Patient number | SOD1-mutation Protein (cDNA) | Zygosity | Diagnosis | Sex | ALS-FRS-R-Score Baseline | ALS-FRS-R-Score tofersen 3 months | Delta ALSFRS-R-Score (tofersen-baseline) |
|---|---|---|---|---|---|---|---|
| **SOD1-ALS patients** | | | | | | | |
| 1 | p.Asp91Ala (c.272A>C) | Hom | ALS | male | 32 | 39 | 7 |
| 2 | p.Asp91Ala (c.272A>C) | Het | ALS | female | 32 | 35 | 3 |
| 3 | p.Asp91Ala (c.272A>C) | Hom | ALS | male | 44 | 44 | 0 |
| 4 | p.Arg116Gly (c.346C>G) | Het | ALS | male | 38 | 33 | -5 |
| 5 | p.Val119_Val12 (c.358-10T) | Het | ALS | female | 34 | 33 | -1 |
| 6 | p.Asp91Val (c.272A>T) | Het | ALS | female | 41 | 35 | -6 |
| 7 | p.Leu145Phe (c.435G>T) | Het | ALS | female | 38 | 37 | -1 |
| 8 | p.Gly94Cys (c.280G>T) | Het | ALS | male | 44 | 43 | -1 |
| 9 | p.Ile114Thr (c.341T>C) | Het | ALS | male | 41 | 39 | -2 |
| **Healthy controls** | | | | | | | |
| - | - | | Tension headache | male | - | - | - |
| - | - | | Facial palsy | female | - | - | - |
| - | - | | Tension headache | male | - | - | - |
| - | - | | Facial palsy | male | - | - | - |
| - | - | | Tension headache | female | - | - | - |
| - | - | | Sinusitis | female | - | - | - |
| - | - | | Tension headache | female | - | - | - |
| - | - | | Idiopathic intracranial hypertension | male | - | - | - |
| - | - | | Unsystematic dizzyness | female | - | - | - |

Hom = homozygous; Het = heterozygous

### Supplementary Methods

#### Participants

Patients were recruited and enrolled from the German MND-NET, a clinical and scientific network of 25 German motor neuron disease centres. We continuously collected data from 9 ALS patients with *SOD1* mutations who participated in the German tofersen early access program (EAP) (Wiesenfarth et al., 2024). All patients were diagnosed with definite laboratory-supported, familial ALS according to the revised El Escorial criteria and were positively tested for a pathogenic *SOD1* variant. The inclusion criteria for participation in the EAP comprised the patients' informed consent as well as the lack of contraindications to a lumbar puncture. Apart from riluzole, recruited patients did not receive any further disease-modifying drug therapy. Patients participating in the long-term extension of the placebo-controlled phase III Tofersen study (VALOR) were excluded from the EAP and this study. The EAP included a dosing phase with intrathecal administration of 100 mg tofersen at day 1, 14 and 28, and a subsequent maintenance phase during which patients received up to 16 doses at intervals of about 28 (at least 21) days. The cohort contains clinical data of 9 patients, which have been published previously by Wiesenfarth et al., 2024.

#### NULISAseq assay

The NULISAseq CNS Disease Panel 120 allows highly sensitive and specific multiplexed analysis of 120 neuro-specific and inflammatory response proteins at the same time. This panel consists of established biomarkers for various neurodegenerative diseases or candidate markers. Many of these proteins have previously not been measured or due to methodological limitations could not be measured in serum or CSF, respectively. NULISAseq assays were performed at Alamar Biosciences as described previously (Feng *et al.,* 2023). Briefly, serum and CSF samples stored at -80$^{o}$C were thawed on ice and centrifuged at 10,000g for 10mins. 10uL supernatant samples were plated in 96-well plates and analysed with Alamar's CNS Disease Panel targeting mostly neuro-degenerative disease related targets as well as inflammation and immune response-related cytokines and chemokines. A Hamilton-based automation instrument was used to perform the NULISAseq workflow, starting with immunocomplex formation with DNA-barcoded capture and detection antibodies, followed by capturing and washing the immunocomplexes on paramagnetic oligo-dT beads, then releasing the immunocomplexes into a low-salt buffer, which were captured and washed on streptavidin beads. Finally, the proximal ends of the DNA strands on each immunocomplex were ligated to generate a DNA reporter molecule containing both target-specific and sample-specific barcodes. DNA reporter molecules were pooled and amplified by PCR, purified and sequenced on Illumina NextSeq 2000.

#### Data analysis

Sequencing data were processed using the NULISAseq algorithm (Alamar Biosciences). The sample- (SMI) and target-specific (TMI) barcodes were quantified, and up to two mismatching bases or one indel and one mismatch were allowed. Intraplate normalisation was performed by dividing the target counts for each sample well by that well's internal control counts. Interplate normalisation was then performed using interplate control (IPC) normalisation, wherein counts were divided by target-specific medians of the three IPC wells on that plate. Data were then rescaled, add 1 and log2 transformed to obtain NULISA Protein Quantification (NPQ) units for downstream statistical analysis.

The transformed datasets were imported into Python (version 4.0.11; JupyterLab) using the pandas package. A Venn diagram was constructed to visualise the commonly expressed proteins in the Serum and CSF using the Matplotlib package. A heatmap with hierarchical clustering based on z-scores was generated for all proteins in the Serum and CSF using Seaborn and Matplotlib packages. Pearson's correlation was performed for the control, baseline, and Tofersen groups separately using the NumPy package, along with further visualisation using heatmaps with hierarchical clustering using the Seaborn and Matplotlib packages. Bar graphs comparing the mean Pearson's correlation coefficients of the healthy controls, baseline, and Tofersen were generated using GraphPad Prism 10. Two-dimensional and three-dimensional PCAs, along with k-means clustering, were performed using the NumPy, Seaborn, Sklearn, and Matplotlib packages for all proteins in serum, all proteins in CSF, and commonly expressed proteins in serum and CSF. Cut-offs for principal components (PCs) were established using the formula 100/ (Total Number of PCs -1). Principal component loadings were extracted from the PCA analysis by creating a new data file using the pandas package and plotted as bar graphs using the matplotlib package. The coordinates of each patient at baseline ($x_1$, $y_1$) and Tofersen ($x_2$, $y_2$) were extracted from PC2 vs. PC3 graphs of all proteins in the serum and CSF. The Euclidean distance was calculated using the following formula: $\sqrt{(x_2{}^2 - x_1{}^2) + (y_2{}^2 - y_1{}^2)}$. Spearman's correlation was performed between ALSFRS-r score and Euclidean distance. Scatterplots to visualise correlations were generated in Python using the seaborn package.

Microsoft Excel and GraphPad Prism (Version 9) were used for data collection and statistical analysis with the following statistical tests: in case of normally distributed data, two independent groups were compared using the unpaired t-test with Welch's correction, in case of non-normal distribution, nonparametric Mann-Whitney test was applied. Tofersen- and baseline-condition were analysed using the paired t-test for normally distributed data, and Wilcoxon matched-pairs signed rank test in case of non-normal distribution comparing samples from similar patients. To evaluate differences among multiple groups, one-way ANOVA followed by the post-hoc Tukey test was used. Statistical significance was set at $p< 0.05$. All data analysed by paired t-test are displayed as mean, including comparison lines for paired data points (patients).

#### Graphical display

Graphical illustrations shown in the figures were adapted from pictures provided by Servier Medical Art (Servier; https://smart.servier.com/) licensed under a Creative Commons Attribution 4.0 Unported License.
